# Global Burden of Stroke Attributable to Kidney Dysfunction, 1992-2021: Age-Period-Cohort Analysis and Projected Trends

**DOI:** 10.1101/2025.04.17.25326046

**Authors:** Chang Li, Xiao Liu, Zhongping Feng, Baofeng Xu, Lina Jin, Rui Liu

## Abstract

**BACKGROUND:** Kidney dysfunction is an important modifiable risk factor for stroke, yet its attributable global burden remains understudied. This analysis quantifies its impact across demographics and projects future trends.

**METHODS AND RESULTS:** Using Global Burden of Disease 2021 data, we analyzed stroke-related deaths and disability-adjusted life years (DALYs) attributable to kidney dysfunction globally, regionally, and nationally, stratified by age, sex, and socio-demographic index (SDI). Trends (1992–2021) were assessed via age-period-cohort (APC) modeling and estimated annual percentage change (EAPC). Contributions of aging, population growth, and epidemiological shifts were quantified through decomposition analysis. Bayesian models projected trends to 2040. Globally, age-standardized mortality rate (ASMR) and age-standardized disability-adjusted life year rate (ASDR) declined (EAPC: −1.85% (95% CI −1.95 to −1.74) and −1.73% (95% CI −1.82 to −1.63)), yet absolute deaths and DALYs rose to 676,000 and 15.009 million in 2021. The burden surged after age 80, disproportionately affecting males and low-SDI regions. Middle-SDI regions showed the steepest declines, while Southern Sub-Saharan Africa experienced rising ASMR and ASDR. Projections suggest continued declines, particularly in females, though disparities persist.

**CONCLUSIONS:** Despite global declines, stroke burden attributable to kidney dysfunction remains elevated in older males and low-SDI regions. Targeted interventions addressing kidney health and equitable healthcare access are critical to mitigating disparities and reducing future burden.

## INTRODUCTION

Globally, stroke continues to rank among the top contributors to disability and death, while kidney dysfunction has gained recognition as an important modifiable risk factor.^1^ Despite advancements in stroke prevention and management, the impact of kidney dysfunction on stroke outcomes has not been uniformly addressed, particularly across different socioeconomic and demographic groups.^2^ The interaction between kidney dysfunction and cerebrovascular disease is complex, involving mechanisms such as hypertension, electrolyte imbalances, inflammation, and endothelial dysfunction.^3^ Uremia can accelerate atherosclerosis by increasing dyslipidemia and induce arterial medial calcification through osteogenic phenotypic changes in vascular smooth muscle cells.^4^ Patients with kidney dysfunction exhibit a greater propensity for thrombosis, with clots differing in structure and function from those in individuals with normal kidney function.^5^ Emerging evidence demonstrates that even subclinical renal impairment, independent of cardiovascular disease or diabetes status, contributes significantly to stroke pathogenesis.^6^ Severe renal impairment demonstrates strong correlations with both clinical progression and functional recovery in embolic stroke of undetermined etiology cases.^7^ Recent ischemic stroke attribution research has classified kidney dysfunction among five key metabolic determinants substantially influencing current and projected ischemic stroke burden.^8^ Renal pathology shows robust associations with silent cerebrovascular lesions, vascular cognitive decline, and dementia, presenting challenges for acute stroke care and long-term prevention strategies.^9^

The past thirty years have witnessed substantial advancements in cerebrovascular disease control, resulting in declining stroke-related morbidity and mortality across multiple populations.^10^ However, the impact of kidney impairment on stroke outcomes has not been uniformly addressed, and disparities in stroke burden persist across different socioeconomic and demographic groups. The GBD initiative establishes a robust epidemiological platform for analyzing disease patterns and associated risk factors, yielding critical data on stroke burden linked to renal pathology across multiple geographic scales. Leveraging GBD 2021 datasets, our investigation quantifies the worldwide impact of kidney dysfunction on stroke from 1992-2021. Using APC methodology, we differentiate the distinct influences of chronological aging, temporal trends, and generational susceptibility on stroke outcomes related to renal dysfunction. Our predictive modeling incorporates Bayesian approaches to forecast stroke trajectories, offering evidence-based projections of renal-related stroke burden. These results advance understanding of preventable stroke determinants and inform targeted strategies for reducing global stroke disparities.

## METHODS

### Study Data

The 2024 GBD 2021 study offers a systematic evaluation of 371 global disease and injury conditions.^1^ Global Health Data Exchange provided stroke burden data (1992-2021) associated with renal impairment across 204 nations, stratified by five SDI tiers: high, high-middle, middle, low-middle, and low development regions. The SDI metric (range 0-1) integrates three development indicators:age-specific fertility rates (<25 years), educational attainment ( ≥ 15 years), time-adjusted per capita income.^11^ Countries were additionally classified into 21 geographical subdivisions. Age stratification comprised 15 categories (25-94 years in 5-year increments plus ≥ 95 years). Kidney dysfunction is generally defined as impaired kidney function due to various causes, resulting in the inability of the kidneys to perform essential physiological functions such as excreting metabolic waste, regulating water-electrolyte balance, and maintaining acid-base balance.^12^ Stroke mortality followed WHO standards, requiring acute-onset focal neurological deficits persisting >24 hours or causing death.^8^

### Statistical Analysis

Age-standardized rates (ASRs) of DALYs and mortality served as key metrics for evaluating temporal patterns in kidney dysfunction-related stroke burden across geographic and demographic strata. These standardized measures enable valid comparisons among populations with varying age distributions or within populations across time periods. We computed EAPC to analyze trends in ASDR and ASMR associated with stroke from 1992 through 2021. We employed log-linear regression modeling of ASR using the equation:

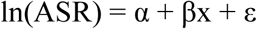

where x denotes calendar year. The EAPC was computed as:

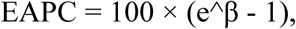

with 95% CI obtained from the regression parameters. Trend interpretation followed these criteria:

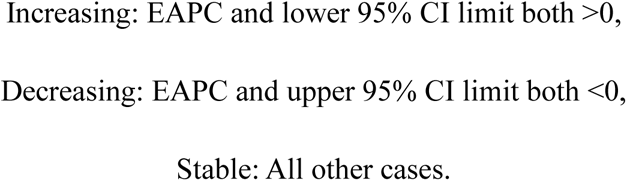

We employed the Das Gupta decomposition method to conduct a decomposition analysis, dissecting changes in DALYs and deaths attributable to stroke caused by kidney dysfunction into three component-level determinants: population age structure, population growth, and epidemiological changes. We quantified the proportional attribution of each determinant to the observed temporal variations.

We applied APC modeling to evaluate temporal disease patterns, specifically examining age, period, and cohort effects. The APC framework requires uniform intervals; thus, we aligned 5-year age brackets with corresponding 5-year calendar periods. Data were structured into 15 five-year age groups (25-29 to ≥95 years), six calendar periods (1992-2016 in 5-year intervals), and 20 derived birth cohorts (1892-1996), enabling full age-period-cohort decomposition while maintaining consistent interval widths as required by APC methodology. Key temporal trends were quantified through net drift (overall annual percentage change reflecting combined period or cohort effects) and local drift (age-specific annual percentage changes). Regional socioeconomic associations were assessed using Spearman’s correlation coefficient between ASDR, ASMR, and SDI across 21 geographical regions. All analyses were conducted using R statistical software (version 4.3.0), with statistical significance set at α=0.05 (two-tailed).

We further applied a Bayesian model to predict age-standardized stroke rates from 2022 to 2040. This model aimed to reveal the distribution of disease across populations and predict future trends in disease burden. The Bayesian model incorporates prior information and sample data to derive posterior distributions, offering greater flexibility and reliability in parameter estimation and prediction.

### Standard Protocol Approvals, Registrations,and Patient Consents

Data used in this study were obtained from the GBD database (https://vizhub.healthdata.org/gbd-results/), an open-source repository that does not contain any personal or identifiable information. As a result, this study was exempt from ethics board review and did not require informed consent from individuals.

### Data Availability

Deidentified datasets generated during this study are available to qualified researchers through reasonable request to the corresponding author, subject to institutional data sharing agreements.

## RESULTS

### Global Trends in Stroke Burden Attributable to Kidney Dysfunction(1992–2021)

From 1992 to 2021, the global number of deaths attributable to stroke caused by kidney dysfunction increased from 481,000 to 676,000. However, the ASMR decreased from 13.18/100,000 (95% UI 8.96/100,000-17.37/100,000) to 8.10/100,000 (95% UI 5.58/100,000-10.73/100,000), with an EAPC of −1.85 (95% CI −1.95- −1.74) (eTable 1, Figure 1 A,C). Similarly, the global number of DALYs attributable to stroke caused by kidney dysfunction increased from 11.09 million to 15.01 million, while the ASDR decreased from 274.96/100,000 (95% UI 200.64/100,000-350.96/100,000) to 174.59/100,000 (95% UI 127.03/100,000-222.85/100,000), with an EAPC of −1.73 (95% CI −1.82- −1.63) (Table 1, Figure 1 B,D). Significant declines in ASMR were observed in high-income regions, including High-income Asia Pacific (EAPC=4.42, 95% CI −4.61- −4.23), Western Europe (EAPC=-4.03, 95% CI −4.19- −3.87), and Australasia (EAPC=-3.96, 95% CI −3.80- −3.58). In contrast, Southern Sub-Saharan Africa experienced an increase in ASMR from 12.70/100,000 (95% UI 8.83/100,000-16.90/100,000) to 14.37/100,000 (95% UI 10.08/100,000-18.83/100,000), with an EAPC of 0.40 (95% CI −0.10- 0.91) (eTable 1, Figure 2 A-B). Similarly, ASDR declined most significantly in High-income Asia Pacific (EAPC=-3.95, 95% CI −4.14- −3.76), Western Europe (EAPC=-3.82, 95% CI −4.00- −3.64), and Australasia (EAPC=-3.53, 95% CI −3.67--3.38), while Southern Sub-Saharan Africa saw an increase in ASDR from 285.75/100,000 (95% UI 211.78/100,000- 365.96/100,000) to 301.93/100,000 (95% UI 226.77/100,000- 386.34/100,000), with an EAPC of 0.18 (95% CI −0.30- 0.66) (Table 1, Figure 2 C-D).

**Figure 1.**
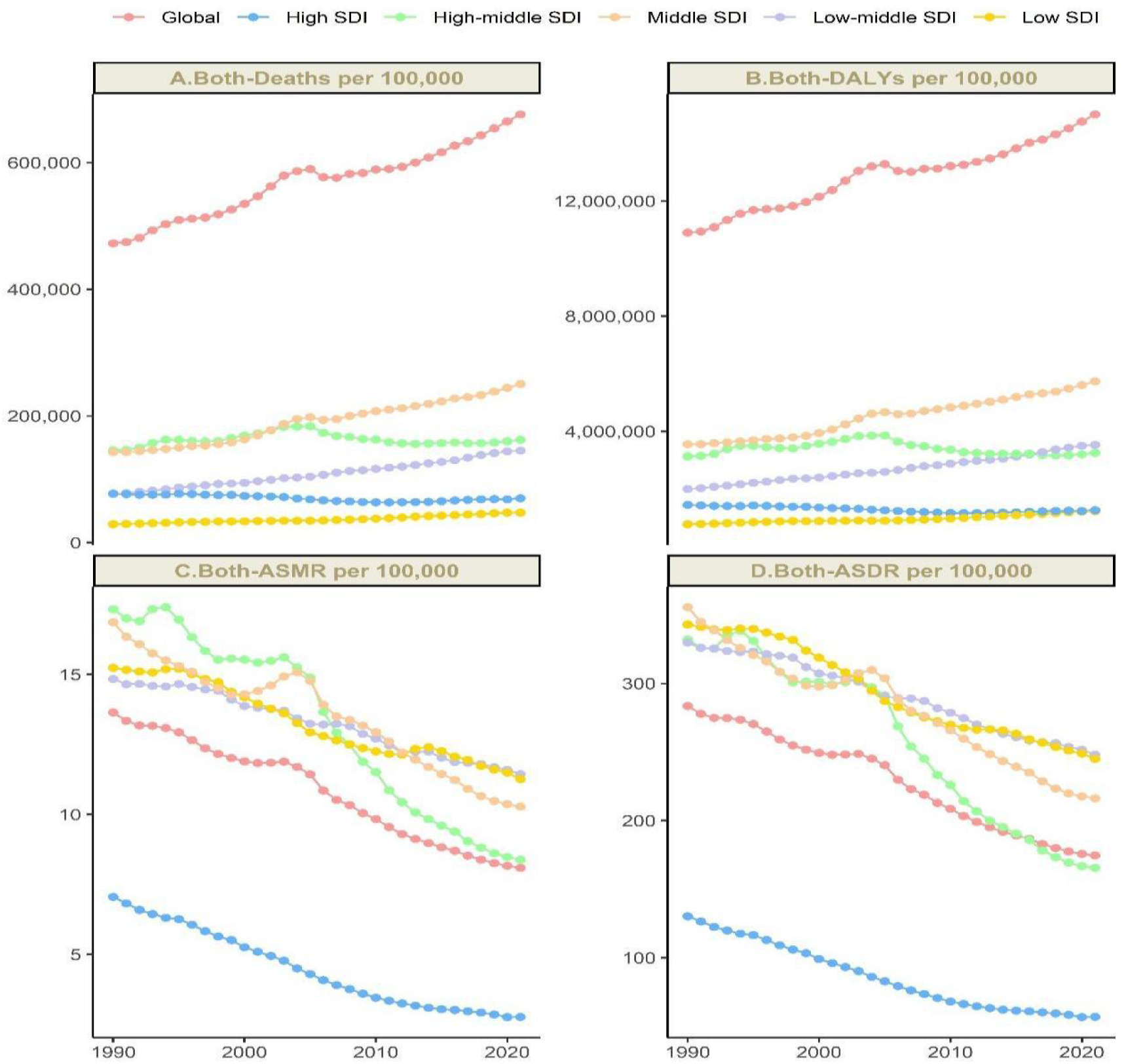
Temporal Trends in Stroke Burden Attributable to Kidney Dysfunction: Deaths(A), DALYs(B), ASMR(C), and ASDR(D) (1992–2021). SDI denotes socio-demographic index; DALYs, disability-adjusted life years; ASMR, age-standardized mortality rate; and ASDR, age-standardized disability-adjusted life-year rate.

**Figure 2.**
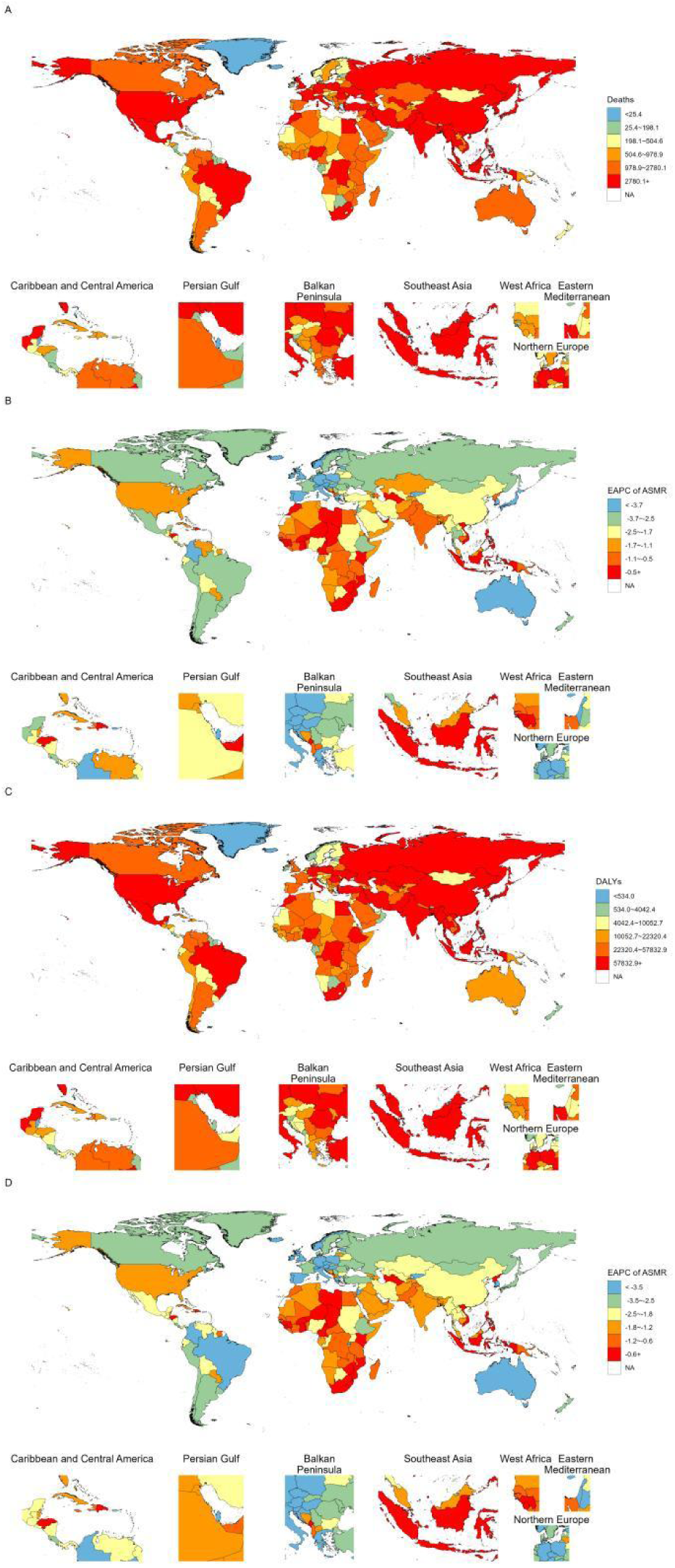
Global Burden of Stroke Attributable to Kidney Dysfunction Across 204 Countries and Territories (2021). (A)Stroke-related deaths due to kidney dysfunction. (B) EAPC in ASMR for kidney dysfunction-related stroke. (C) DALYs due to kidney dysfunction-related stroke. (D) EAPC in ASDR for kidney dysfunction-related stroke. EAPC denotes estimated annual percentage change; DALYs, disability-adjusted life years; ASMR, age-standardized mortality rate; and ASDR, age-standardized disability-adjusted life-year rate.

**Table 1.**
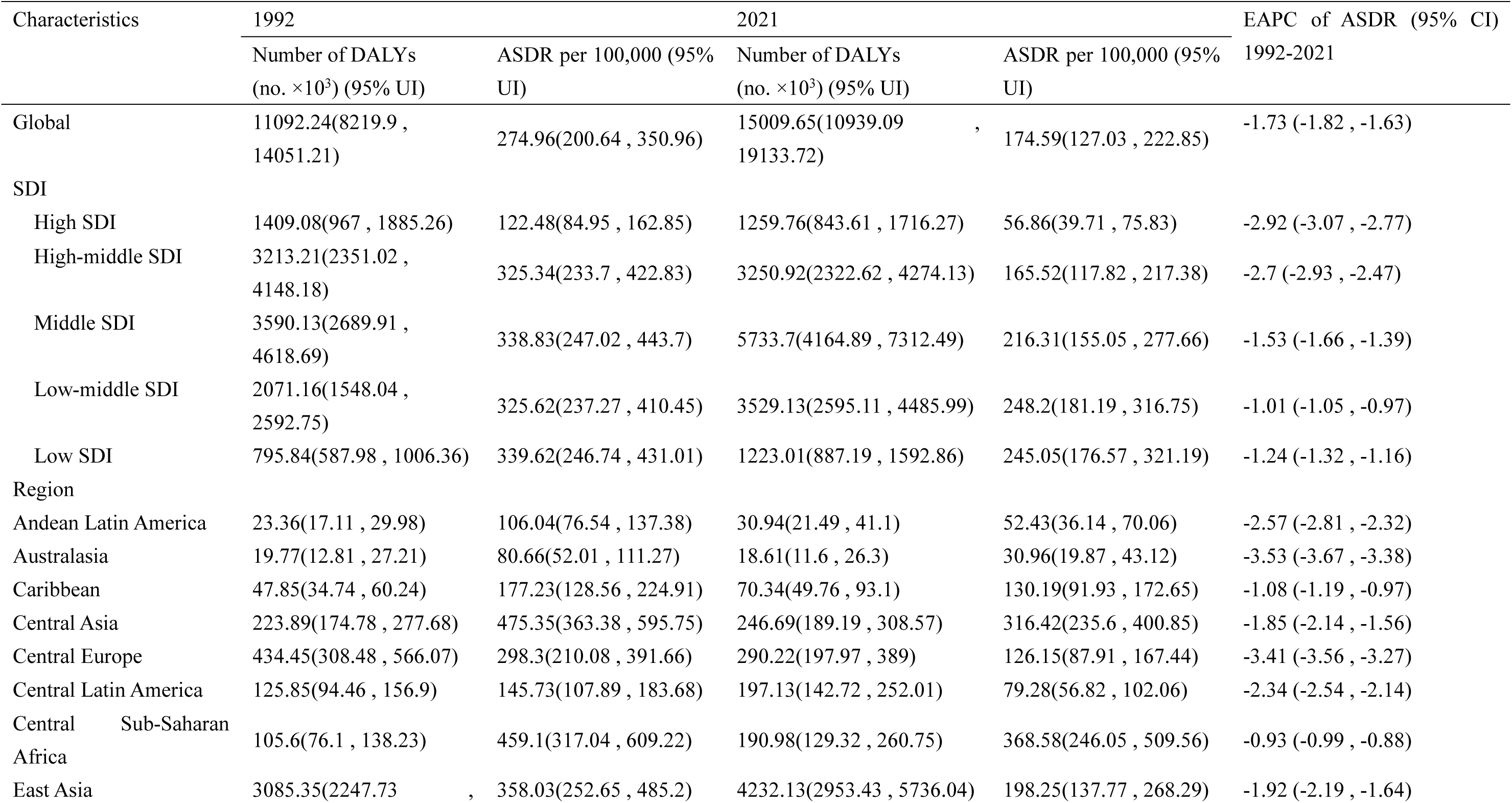
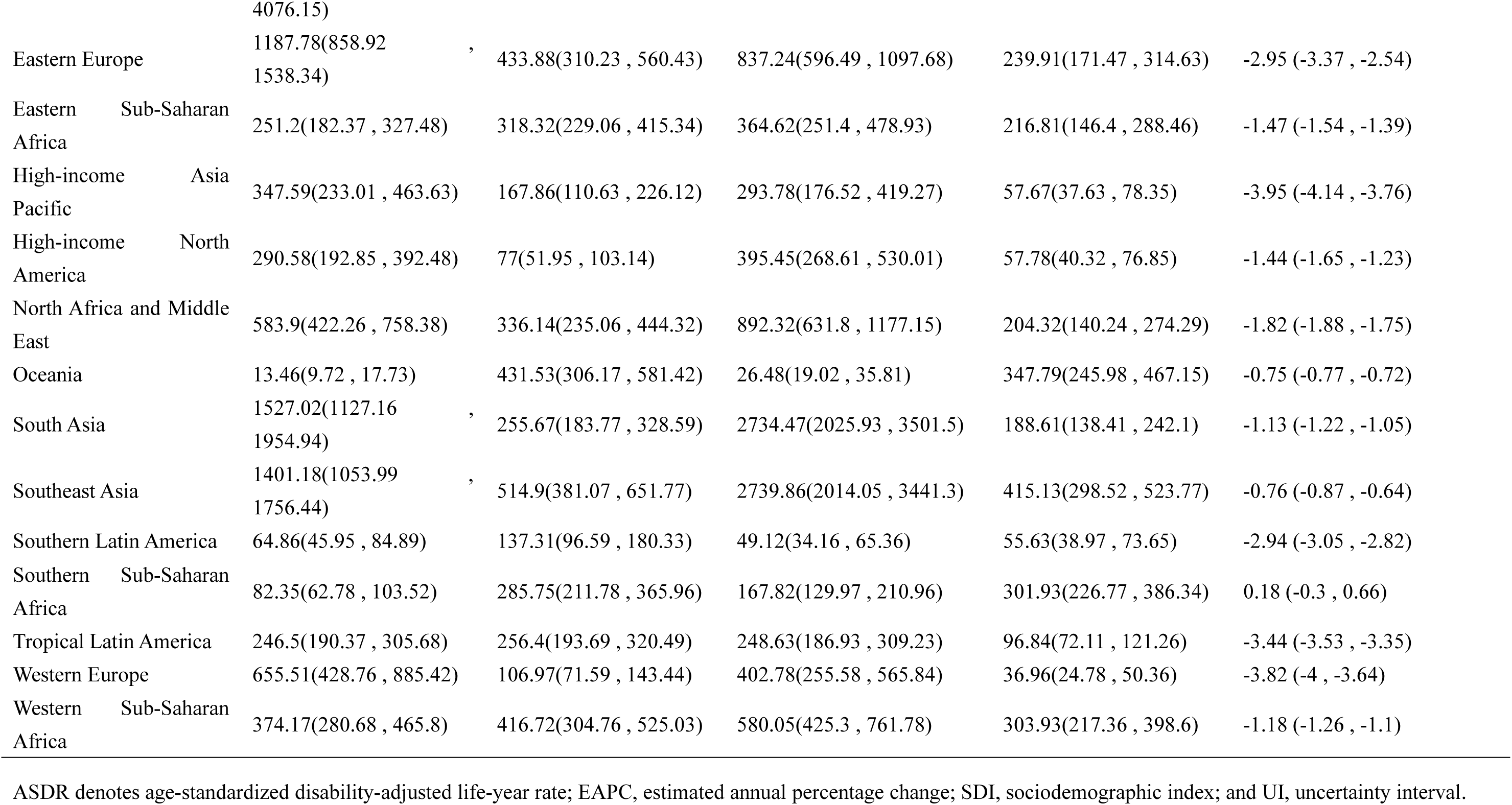
DALYs and ASDR for Stroke Attributable to Kidney Dysfunction in 1992 and 2021, with Temporal Trends (1992–2021).

### Age, Sex, and SDI in Stroke Burden (2021)

The 2021 ASDR for stroke related to kidney dysfunction demonstrated a progressive age-dependent increase, attaining maximal values in nonagenarians (≥95 years). DALYs exhibited distinct sex-specific trajectories: while peaking earlier in males (65-69 years), the maximal burden occurred in females aged 70-74 years, followed by subsequent declines in older age strata for both sexes. Males exhibited higher DALYs than females from ages 25–29 to 70–74, but beyond this age range, females had higher DALYs. Cases were relatively low in both sexes under 30 years and over 95 years of age. The greatest DALYs proportions occurred in middle-high to low-middle SDI regions, particularly affecting adults aged 50-89 years, with peak burdens observed in these middle-development regions(Figure 3).

**Figure 3.**
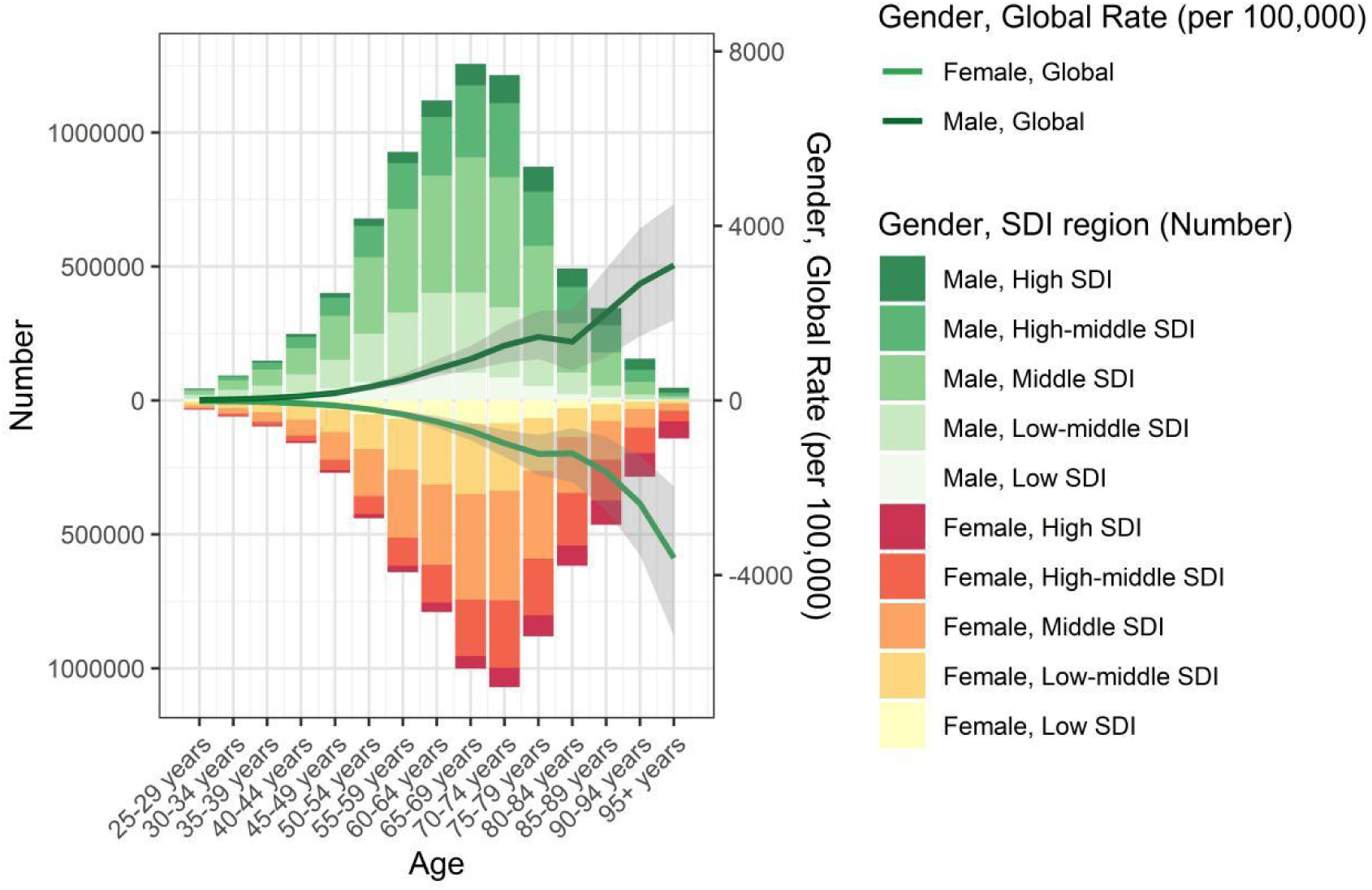
Age-, Sex-, and SDI-Specific Trends in DALYs for Stroke Attributable to Kidney Dysfunction (2021). SDI denotes socio-demographic index; and DALYs, disability-adjusted life years.

### Decomposition Analysis

This study incorporated demographic parameters (population size, age distribution) and standardized mortality metrics to assess disease burden trends. Globally, kidney dysfunction-related stroke mortality and DALYs showed marked increases, with the most substantial burden concentrated in middle-to-low SDI regions. Population expansion constituted a key determinant of these observed epidemiological shifts, particularly in developing regions. Epidemiological changes exerted a protective effect globally and across all five SDI regions. Specifically, in middle, low-middle, and low SDI regions, population growth and aging accelerated this increase, while their impact was less pronounced in other regions. Aging and population growth emerged as critical factors driving the escalating burden of stroke attributable to kidney dysfunction (eTable 2, Figure S1).

### Temporal Trends in Stroke-Related DALYs Across Age Groups

Figure 4A and eTable 3 present the net drift and local drift calculated using the APC model for global and different SDI regions. The net drift is depicted by colored horizontal lines, while local drift is represented by curved solid lines. A black horizontal dashed line marks the reference point where the EAPC=0. The colored horizontal solid lines, along with their associated ranges, indicate the net drift and corresponding 95% CIs for each group. The net drift for both female and male groups globally was less than 0, indicating an overall declining trend in ASDR for stroke attributable to kidney dysfunction. For females, the declining trend weakened from ages 25–29 to 50–54 and 75–79 and above, while it strengthened from ages 55–59 to 65–69. For males, the declining trend weakened from ages 25–29 to 55–59 and strengthened for those aged 60 and above. In high-SDI regions, the local drift curve for males crossed the 0 line at two points, indicating a positive trend for individuals aged 40–49 (EAPC > 0) and a negative trend for those aged 25–39 and 50 years and older (EAPC < 0). This suggests an increasing burden among males aged 40–49, while a decline was observed in other age groups. In contrast, the local drift curve for females in high-SDI regions remained entirely below zero, reflecting a consistent downward trend. Patterns in middle-high and low-middle SDI regions aligned with global trends. In middle-SDI regions, the decline in burden became more pronounced over time, whereas in low-SDI regions, the rate of decline progressively weakened.

**Figure 4.**
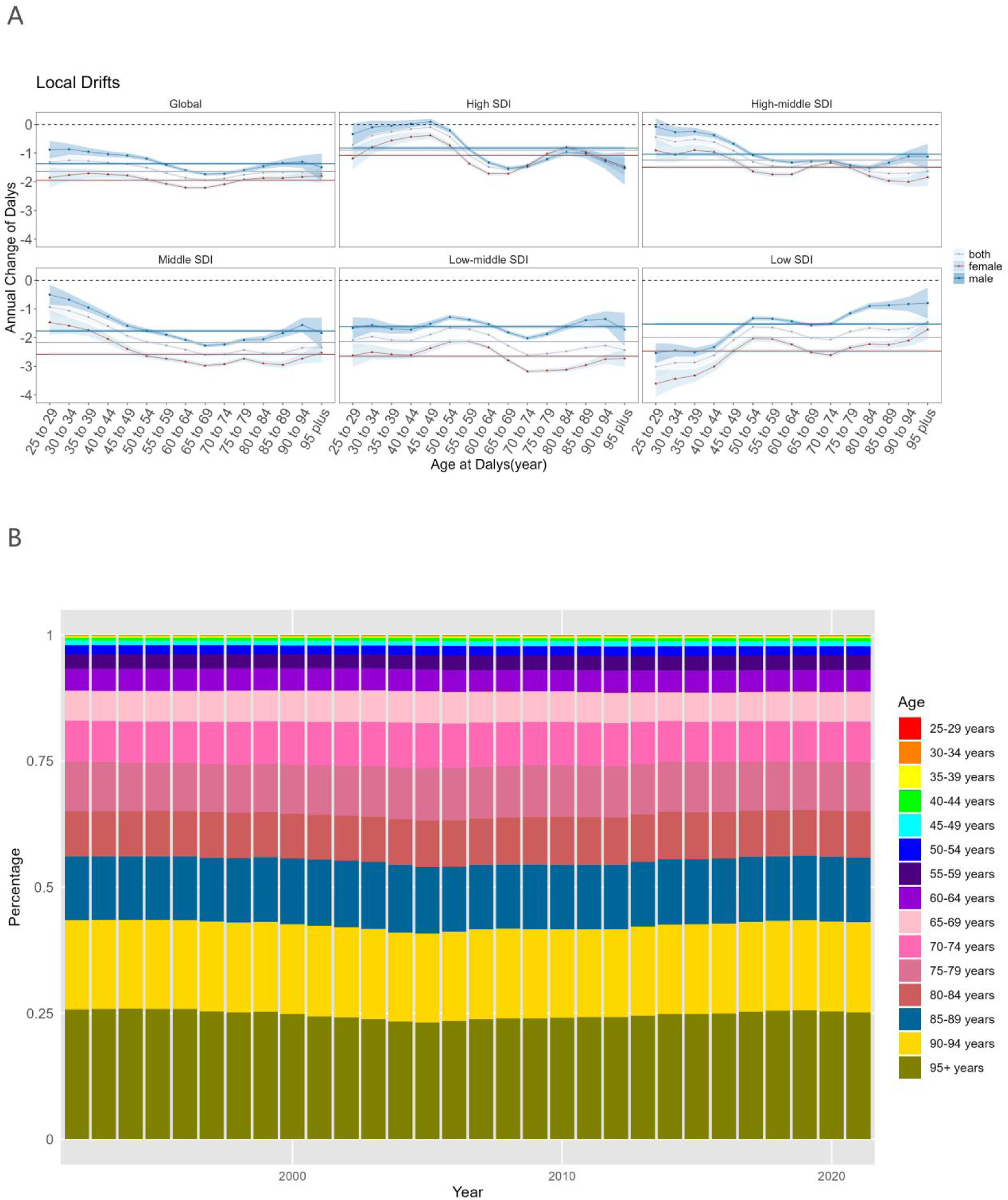
Temporal Trends and Age-Specific Distribution of DALYs for Stroke Attributable to Kidney Dysfunction by SDI Quintiles (1992–2021). (A) Local drift in DALYs for kidney dysfunction-related stroke (1992–2021). Dots and shaded areas represent estimates with 95% CIs. (B) Age-specific distribution for kidney dysfunction-related stroke (1992–2021). DALYs denote disability-adjusted life years.

Figure 4B demonstrates the temporal shifts in the age-specific distribution of DALYs associated with stroke due to kidney dysfunction between 1992 and 2021. The risk of stroke due to kidney dysfunction increased progressively with age, with higher proportions observed in older age groups. Globally, younger age groups (25–49 years) contributed minimally to the burden, whereas middle-aged and older groups (50–74 years) showed a marked increase in proportion. The oldest age group (≥75 years) accounted for the highest proportion, surpassing 75% of the total burden. These findings highlight the predominance of older age groups in driving the burden over the three-decade period. No significant annual variations in age-specific proportions were observed from 1992 to 2021.

### APC Analysis

Figure 5A-C presents the APC analysis of DALYs associated with stroke resulting from kidney dysfunction, demonstrating distinct temporal patterns across these demographic dimensions. Globally, DALYs increased gradually with age, rising sharply after 80–84 years and peaking in the 95 years and older age group. Both sexes exhibited comparable trends, though males demonstrated marginally higher DALYs than females. Within high-SDI regions, age-specific DALYs exhibited a bimodal distribution: an initial peak at 70-74 years followed by a transient decline, with subsequent resurgence in the oldest age groups (≥80-84 years). Trends in other SDI regions aligned with global patterns, with low-middle and low-SDI regions displaying a greater burden among females compared to males, particularly in the age group of 80–85 years and older (Figure 5A).

**Figure 5.**
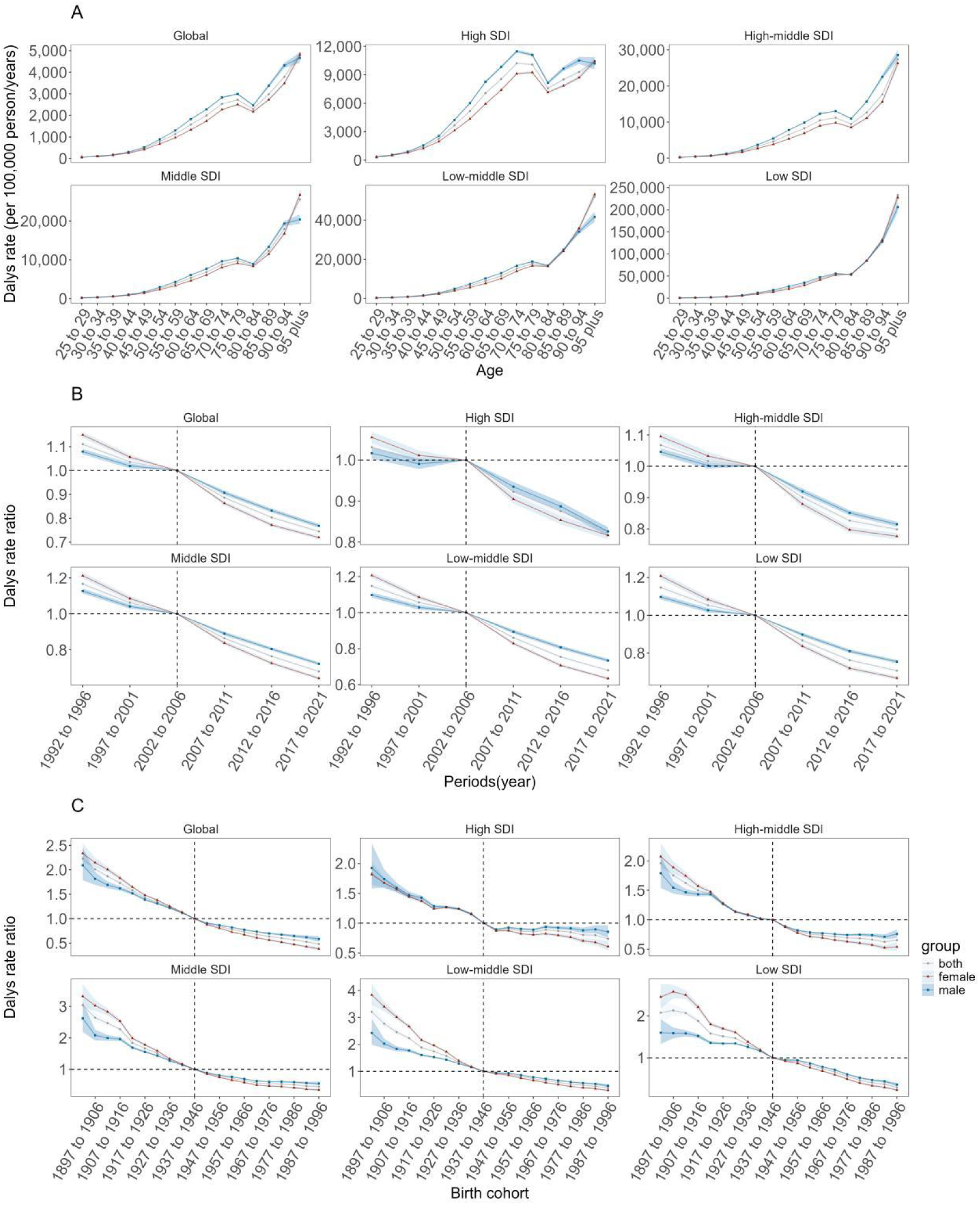
APC Effects on DALYs from 1992 to 2021 for Stroke Attributable to Kidney Dysfunction. (A) Age effects on DALYs for kidney dysfunction-related stroke (1992–2021). (B) Period effects on DALYs for kidney dysfunction-related stroke (1992–2021). (C) Cohort effects on DALYs for kidney dysfunction-related stroke (1992–2021). DALYs denote disability-adjusted life years.

In the period cohort, from 1992 to 2021, stroke attributable to kidney dysfunction declined for both males and females. Females exhibited higher relative risks before the 2002–2006 cohort, but their decline was faster than that of males thereafter. The global period effect demonstrated consistent attenuation, particularly marked in middle-SDI regions where the steepest reduction was observed (Figure 5B). Regarding the birth cohort effect, trends were similar to the period effect. Sex disparities were most evident in high-SDI areas, with males showing significantly elevated risks compared to females. Conversely, low-SDI regions displayed peak risk in the 1897-1906 birth cohort, followed by progressive risk diminution (Figure 5C).

### Future Forecasts of Global Burden

Global projections of stroke-related deaths and DALYs attributable to kidney dysfunction were estimated for the period 2022–2040, stratified by sex. A decline in both deaths and DALYs is anticipated across the overall population, as well as within male and female subgroups. Significant sex disparities were observed, with females projected to have lower numbers of deaths and DALYs compared to males by 2024, and a steeper decline in trends than males (Figure 6).

**Figure 6.**
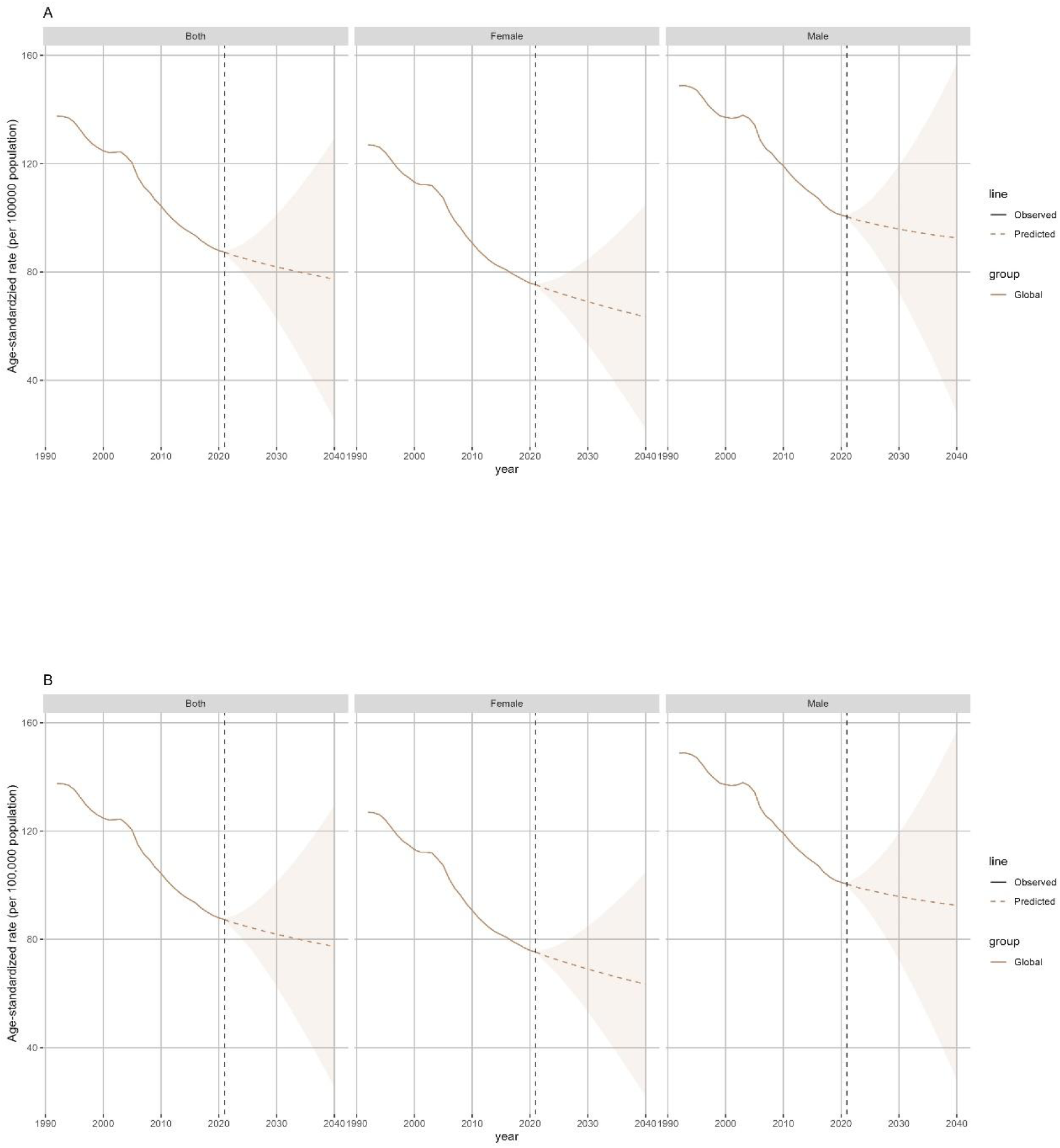
Future Forecasts of Deaths(A) and DALYs(B) for Stroke Caused by Kidney Dysfunction.

### Correlation Between ASDR, ASMR, and SDI

Efigure 2 depicts the association between ASDR for stroke attributable to kidney dysfunction and the SDI across 21 global regions, as categorized by the GBD study (1992–2021). Colored lines indicate temporal trends for individual regions, with each data point representing a specific year within the study period. R = −0.416 indicates a negative correlation between SDI and ASDR, with a moderate strength of association, suggesting that ASDR decreases as SDI values increase. Figure S7 illustrates the relationship between ASMR attributable to kidney dysfunction-related stroke and SDI across 21 global regions classified by GBD from 1992 to 2021. R = −0.363 reflects a weak negative correlation between SDI and ASMR, indicating that ASMR declines with increasing SDI values.

## DISCUSSION

Our investigation provides a comprehensive assessment of stroke burden associated with kidney dysfunction across multiple geographic scales (global, regional, national) during 1992-2021. Analysis revealed significant disparities in stroke resulting from kidney dysfunction epidemiology by SDI stratification and sex, with thirty-year temporal trends and projections extending to 2040. This trend may be attributed to improvements in healthcare, advancements in medical treatments, and better management of risk factors such as hypertension and diabetes, which are closely linked to both stroke and kidney dysfunction.^13,14^ Mortality attributable to stroke mediated by kidney dysfunction demonstrated a consistent rise, rising from 481,000 in 1992 to 676,000 in 2021, while the ASMR decreased from 13.18/100,000 (95% UI 8.96/100,000 to 17.37/100,000) to 8.10/100,000 (95% UI 5.58/100,000 to 10.73/100,000), with an EAPC of −1.85 (95% CI −1.95 to −1.74). The reduction in ASDR reflects advancements in stroke prevention, early detection, and treatment, particularly in high-income regions with stronger healthcare systems.^15^ Notable disparities were observed across the 21 GBD regions. The most significant declines in ASDR occurred in High-income Asia Pacific, Western Europe, and Australasia, with EAPCs of −3.95 (95% CI −4.14 to −3.76), −3.82 (95% CI −4.00 to −3.64), and −3.53 (95% CI −3.67 to −3.38), respectively. These regions likely benefited from well-established healthcare systems, early detection, and effective management of kidney dysfunction and its complications.^16^ Conversely, Southern Sub-Saharan Africa saw an increase in ASDR, from 285.75/100,000 (95% UI 211.78/100,000 to 365.96/100,000) to 301.93/100,000 (95% UI 226.77/100,000 to 386.34/100,000), with an EAPC of 0.18 (95% CI −0.30 to 0.66). Several contributory factors may explain this increase, including suboptimal healthcare access, poorly managed comorbidities, and rising incidence of kidney dysfunction across the region.^17^

Our analysis reveals marked inequalities in stroke burden related to kidney dysfunction when stratified by sex, age categories, and SDI tiers. In 2021, the proportion of DALYs was higher among males than females, reflecting gender differences in disability caused by stroke related to kidney dysfunction.^18^ The impact of stroke consequent to kidney dysfunction exhibited an age-dependent progression, with the most severe manifestations occurring in patients over 80 years old, demonstrating the amplified risks associated with advanced age and declining renal function.^19^ When examining SDI-stratified populations, the middle-development spectrum (encompassing middle-high through low-middle SDI areas) demonstrated peak DALYs concentrations among adults aged 50-89 years. This finding suggests that despite progress in reducing the stroke burden in these regions, older populations remain at significant risk.^20^ In sharp contrast, low-SDI regions - particularly throughout Sub-Saharan Africa - are experiencing growing cerebrovascular morbidity associated with renal pathology, largely attributable to constrained medical infrastructure and escalating rates of chronic kidney disease.^21^

Our decomposition analysis highlights the substantial contribution of demographic shifts, particularly population expansion and aging populations, to increasing kidney dysfunction-related stroke burden, with most pronounced effects in middle-to-low SDI regions. These results align with established literature documenting how decelerating population growth and aging demographics shape global disease burden patterns.^22^ Rising global life expectancy has positioned age-related neurological conditions, including stroke and dementia, among the leading contributors to mortality and disability.^23^ Rapid population growth in low-income countries has been shown to exacerbate the stroke burden, highlighting the urgent need for enhanced healthcare infrastructure.^24^ The observed protective influence of epidemiological transitions likely reflects advancements in controlling major vascular risk factors, including hypertension and diabetes mellitus, as evidenced in previous studies.^25^

APC analysis offers a nuanced understanding of temporal trends in stroke burden attributable to kidney dysfunction. Globally, both males and females exhibited negative net drift, indicating a declining trend in ASDR. However, weaker declines in younger (25–54 years) and older (75+ years) age groups suggest that these populations may require targeted interventions.^26^ For females, the decline in ASDR weakened between ages 25–29 and 50–54, as well as after 75–79 years, while it strengthened between 55–59 and 65–69 years. This pattern may reflect differences in healthcare utilization and risk factor management across age groups. Younger women may be less likely to engage in regular medical care for chronic conditions like hypertension or diabetes, while older women often encounter healthcare access barriers due to mobility limitations or comorbidities.^27^ For males, the decline in ASDR weakened between ages 25–29 and 55–59 but strengthened after 60 years. This demographic pattern likely stems from elevated rates of modifiable risk factors (tobacco use, excessive alcohol intake, and adiposity) in middle-aged male populations, which may offset benefits from enhanced medical care availability and therapeutic advances. In high-SDI regions, local drift for males indicated an increase in ASDR among the 40–49 age group, whereas declines were observed in other age groups.^28^ This finding suggests that middle-aged men in high-income regions may face a unique burden due to lifestyle factors or delayed diagnosis of kidney dysfunction.^29^ In low-middle SDI regions, trends largely mirrored global patterns but with varying magnitudes of decline. Middle-SDI regions showed a gradual strengthening of ASDR declines, likely due to improvements in healthcare infrastructure and increased awareness of kidney dysfunction and stroke prevention.^30^ Low-SDI regions displayed attenuated mortality declines, mirroring systemic barriers to healthcare delivery and suboptimal control of cardiovascular risk factors.^31^ Temporal analyses revealed substantial period and cohort variations, highlighting the cumulative benefits of evolving treatment paradigms and population health initiatives across generations. The consistent downward trends in period and cohort effects, particularly in middle-SDI regions, highlight the critical role of sustained investments in healthcare systems and preventive strategies.^32^ Persistent disparities in low-SDI areas accentuate the urgent requirement for precision public health strategies to bridge gaps in medical infrastructure and service quality.

Future projections (2022-2040) suggest sustained decreases in renal-related stroke mortality and DALYs, with more pronounced improvements anticipated in female cohorts relative to males. This trend may be attributed to better management of kidney dysfunction and stroke risk factors among women, as well as differences in healthcare-seeking behavior.^33^ Nevertheless, persistent gender disparities in stroke burden highlight the need for gender-specific interventions.^34,35^

The negative correlations between SDI and both ASDR and ASMR (R = −0.416, R = −0.363) suggest that higher SDI regions tend to experience a lower stroke burden. This observation aligns with existing evidence on the social determinants of health, highlighting the impact of socioeconomic development on mitigating the burden of non-communicable diseases.^1^ However, the moderate and weak correlations also suggest that other factors, such as healthcare quality, public health policies, and cultural factors, play significant roles in influencing stroke burden.^36^

Our study has several notable limitations. As the analysis used modeled GBD data, it relies on the accuracy and validity of the methods and statistical modeling approaches employed. While the GBD study employs advanced methodologies to address missing data and enhance accuracy, variations in data availability and quality may still affect the results. Additionally, our classification of kidney dysfunction follows established GBD definitions, which may not fully align with clinical standards. Finally, while APC analysis provides valuable insights into temporal trends, its reliance on population-level data presents inherent limitations.

## CONCLUSIONS

Our longitudinal assessment demonstrates complex, evolving trajectories in nephrogenic stroke epidemiology between 1992-2021, reflecting dynamic interactions between medical, demographic, and socioeconomic factors. Although global trends show declines in age-standardized rates, the absolute burden has increased, particularly in low-SDI regions. Significant disparities exist across sex, age, and regions, with older populations and males bearing a disproportionate burden. These evidence-based findings mandate tailored intervention frameworks to mitigate growing stroke burdens in medically underserved populations and disadvantaged geographic regions. The projected decline in stroke burden over the next two decades is encouraging but must be interpreted cautiously. Sustained investment in healthcare infrastructure, public health initiatives, and research is essential to maintain these gains and address persistent disparities. Policymakers and healthcare providers must prioritize equitable access to care and tailored interventions to ensure that the benefits of medical advancements reach all populations.

## Non-standard Abbreviations and Acronyms

APC: age-period-cohort
ASDR: age-standardized disability-adjusted life year rate
ASMR: age-standardized mortality rate
ASR: age-standardized rates
DALYs: disability-adjusted life years
EAPC: estimated annual percentage change
GBD: Global Burden of Disease
SDI: socio-demographic index

## Acknowledgments

We sincerely thank the Institute for Health Metrics and Evaluation, United Nations, and World Bank for providing open-access data essential to this study, and acknowledge helpful discussions with colleagues.

## Supplemental Material

Tables S1–S3

Figures S1–S2

